# Evaluation of Facial Protection Against Close-Contact Droplet Transmission

**DOI:** 10.1101/2021.02.09.21251443

**Authors:** Thomas B. Stephenson, Courtney Cumberland, Geoff Kibble, Christopher Church, Sheila Nogueira-Prewitt, Sebastian MacNamara, Delbert A. Harnish, Brian K. Heimbuch

**Author notes:** Corresponding Author: Delbert Harnish, 430 W. 5^th^ Street, Panama City, FL 32401. Funding: Occupational Safety and Health Administration (OSHA) under Prime contract #1605DC□19□A□0010 with Project Enhancement Corporation (PEC) and Task Order #ARA 20-775-01 with Applied Research Associates.

## Abstract

**Background:** Face shields are used as an alternative to facemasks, but their effectiveness in mitigating the spread of SARS-CoV-2 is unclear. The goal of this study is to compare the performance of face shields, surgical facemasks, and cloth facemasks for mitigation of droplet transmission during close contact conditions.

**Methods:** A novel test system was developed to simulate droplet transmission during close contact conditions using two breathing headforms (transmitter and receiver) placed 4 feet apart with one producing droplets containing a DNA marker. Sampling coupons were placed throughout the test setup and subsequently analyzed for presence of DNA marker using quantitative PCR.

**Results:** All PPE donned on the transmitter headform provided a significant reduction in transmission of DNA marker to the receiver headform: cloth facemask (78.5%), surgical facemask (89.4%), and face shield (96.1%). All PPE resulted in increased contamination of the eye region of the transmitter headform (9,525.4% average for facemasks and 765.8% for the face shield). Only the face shield increased contamination of the neck region (207.4%), with the cloth facemask and surgical facemask resulting in reductions of 85.9% and 90.2%, respectively.

**Conclusions:** This study demonstrates face shields can provide similar levels of protection against direct droplet exposure compared to surgical and cloth masks. However, all PPE tested resulted in release of particles that contaminated surfaces. Contamination caused by deflection of the user’s exhalation prompts concerns for contact transmission via surfaces in exhalation flow path (e.g., face, eyeglasses, etc.).

## Introduction

The COVID-19 pandemic has elevated attention for facial protection to a level previously unseen in the United States. American workers in many occupational settings are required to wear face coverings to mitigate virus transmission by asymptomatic and pre-symptomatic individuals. SARS-CoV-2, the causative agent for COVID-19, is primarily transmitted via respiratory droplets.^1^ While the Centers for Disease Control and Prevention (CDC) has provided guidance that facemasks are effective for source control and droplet protection,^2^ the utility of face shields is still in question.^3^

Studies to assess the effectiveness of different types of face coverings tend to focus on cough or sneeze simulations that produce high velocity droplets and obtain measurements close to the mask after a short duration.^4,5,6^ Such simulated coughing studies have demonstrated face shields can reduce user exposure to droplets by 96%, but do not contain all aerosols expelled by the user.^7,8^ However, it remains unclear how face shields and other types of face coverings perform under normal respiration conditions, which better represent asymptomatic/pre-symptomatic spreaders of SARS-CoV-2. In these cases, transmission can occur via droplets produced through normal respiration and/or conversation during close contact conditions, which the CDC defines as exposure to a symptomatic, asymptomatic, or pre-symptomatic carrier at a distance of less than 6 feet for at least 15 minutes over a 24 hour period.^9^ Thus, it is critical to simulate close contact conditions to fully understand the effectiveness of face shields and other face coverings at preventing droplet spread from a carrier to other individuals.

Simulating normal respiratory output requires that both droplet size distribution and concentration adequately represent conditions produced by humans. Though characterization of respiratory secretions has been studied for almost 80 years, the results of these studies are wide-ranging and can be influenced by a number of factors.^10,11^ Peer-reviewed literature generally supports a droplet size distribution for respiratory secretions ranging from 10 – 100 µm in size.^12,13^ The number of droplets produced during respiration is another critical factor. Xie *et al*. quantifies total droplet mass output to be ∼0.08 g when speaking for ∼1 minute, which would yield approximately 1.2 g over 15 minutes.^12^

The objective of this study was to assess the performance of three types of PPE – cloth facemask, surgical facemask, and face shield – for their ability to mitigate droplet transmission during close contact transmission conditions as defined by the CDC. The intended droplet size distribution will range from 10 – 100 µm in size and volume output will target 0.08 g/min during low work rate respiratory conditions. The results of this study will be used to inform the general public regarding effectiveness of different PPE types against droplet transmission.

## Methods

### Test Samples

Three types of PPE were evaluated during this study: cloth facemasks (RN15763, Hanes, Winston-Salem, NC); surgical facemasks (B087YPTXT3, Alertcare, San Francisco, CA); and face shields (PPE USA, Hunt Valley, MD).

### Test System

The experimental setup consisted of two ISO Medium 3D-printed plastic headforms,^14^ with one headform producing droplets (transmitter) expelled towards the other headform (receiver) to simulate droplet transmission (Figure 1). Each headform has a 1-in orifice at the mouth through which breathing occurred. The transmitter headform was connected to a breathing machine designed to inhale/exhale using a low work rate (minute ventilation = 25 liters per minute, breathing rate = 21 breaths per minute, tidal volume = 1.2 L).^15^ En route to the headform, the air passed through 1-in diameter flexible PVC tubing into a cylindrical spray chamber measuring 15 cm in diameter and 70 cm in length. The spray chamber was fitted with a full-cone misting nozzle (3178K62, McMaster-Carr, Atlanta, GA) having an orifice of 0.025 cm and an 80° spray angle to introduce droplets into the spray chamber. The misting nozzle was connected to a valved tank pressurized to 60 psi and pre-filled with 0.3% mucin solution inoculated with a DNA marker. Air flow exited the spray chamber through the mouth orifice during exhalation only. A Spraytec droplet sizer (Malvern Panalytical, Westborough, MA) was positioned at the orifice of the transmitter to measure the droplet size distribution. The receiver headform was connected to a QuickLung BREATHER™ System (IngMar Medical, Pittsburgh, PA) operating in the Eupnea mode (24 breaths per minute, 720 mL tidal volume).

**Figure 1.**
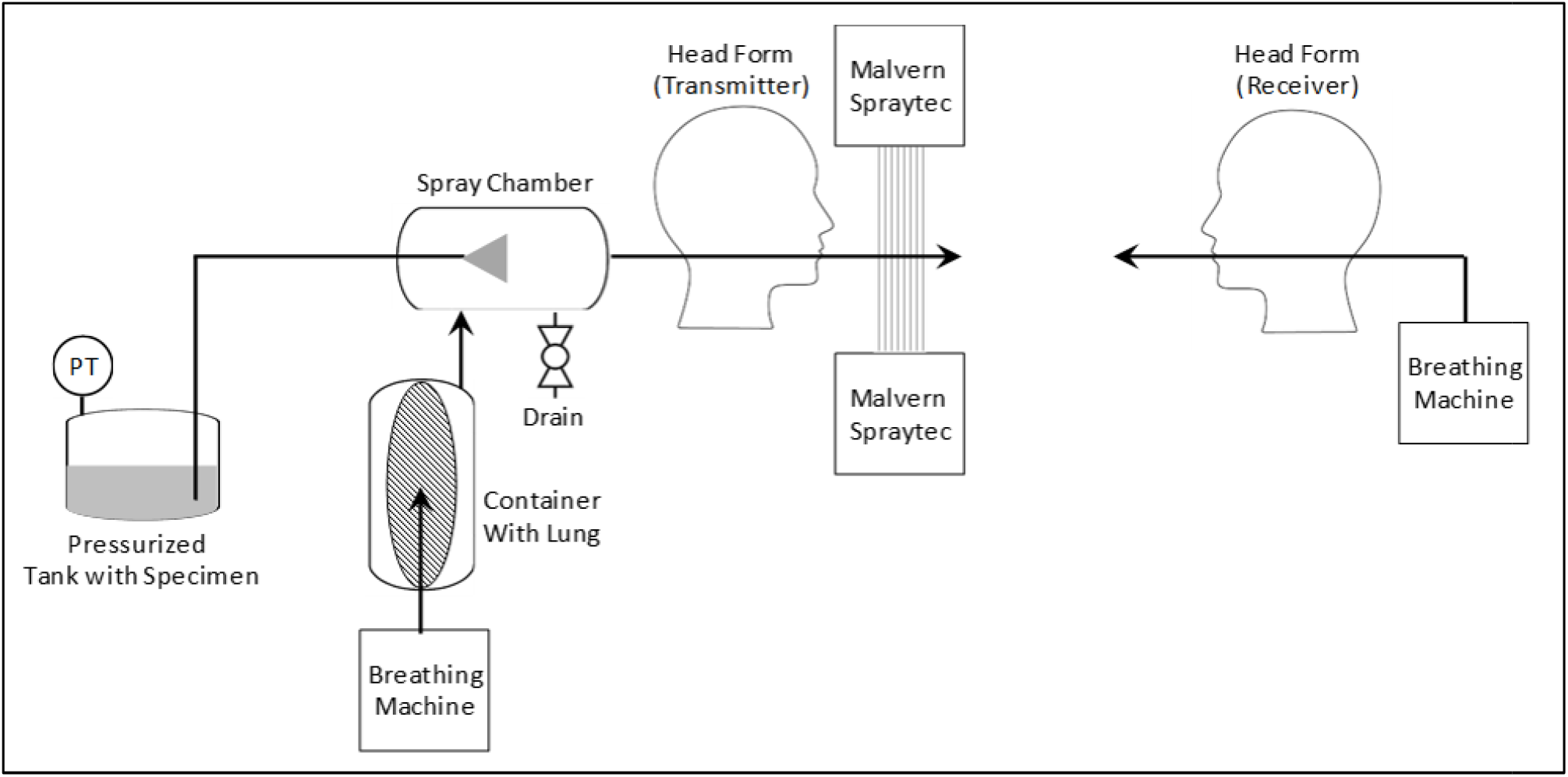
Droplet Transmission Test System Overview.

### Test Analyte

The DNA marker used for this study was a 111-bp DNA fragment (λ-111) amplified from the lambda bacteriophage (λ phage) genome using a previously published qPCR primer set.^16^**Error! Bookmark not defined**. λ-111 was purified using a DNA Clean and Concentrator-100 kit (D4029, Zymogen) and the identity of the DNA fragment was confirmed using gel electrophoresis. DNA concentration was determined using a Synergy LX multi-mode plate reader with a Take3 plate (Biotek, Winooski, VT).

### Droplet Transmission Tests

Both headforms were positioned in a non-ventilated room (∼11’ × 11’ × 10’) lined with anti-static polyethylene film (ASFR612, Americover, Escondido, CA). Sampling coupons were circular, 2.5-cm diameter, 0.4-µm polycarbonate filters (PCT0425100; Sterlitech) on top of a 3.2-cm diameter diameter cellulose filter paper backing (CFP1-032; Sterlitech, Kent, WA). Coupons were placed at 1, 2, 3, and 4 feet directly in front of the transmitter headform (∼18.5” below the headform mouth) in addition to the headform locations (Figure 2). Coupons were placed onto the headforms using double-sided tape, and metal pins were used to hold the coupons to the cellulose backing when needed; coupons placed between and behind headforms were each held in an aluminum weighing dish. Four PPE conditions were tested: 1) Cloth facemask on the transmitter headform, 2) Surgical facemask on the transmitter headform, 3) Face shield on the transmitter headform, and 4) no PPE on the transmitter headform (positive control). For all tests, no PPE was donned onto the receiver headform. Three replicates were performed for each condition.

**Figure 2.**
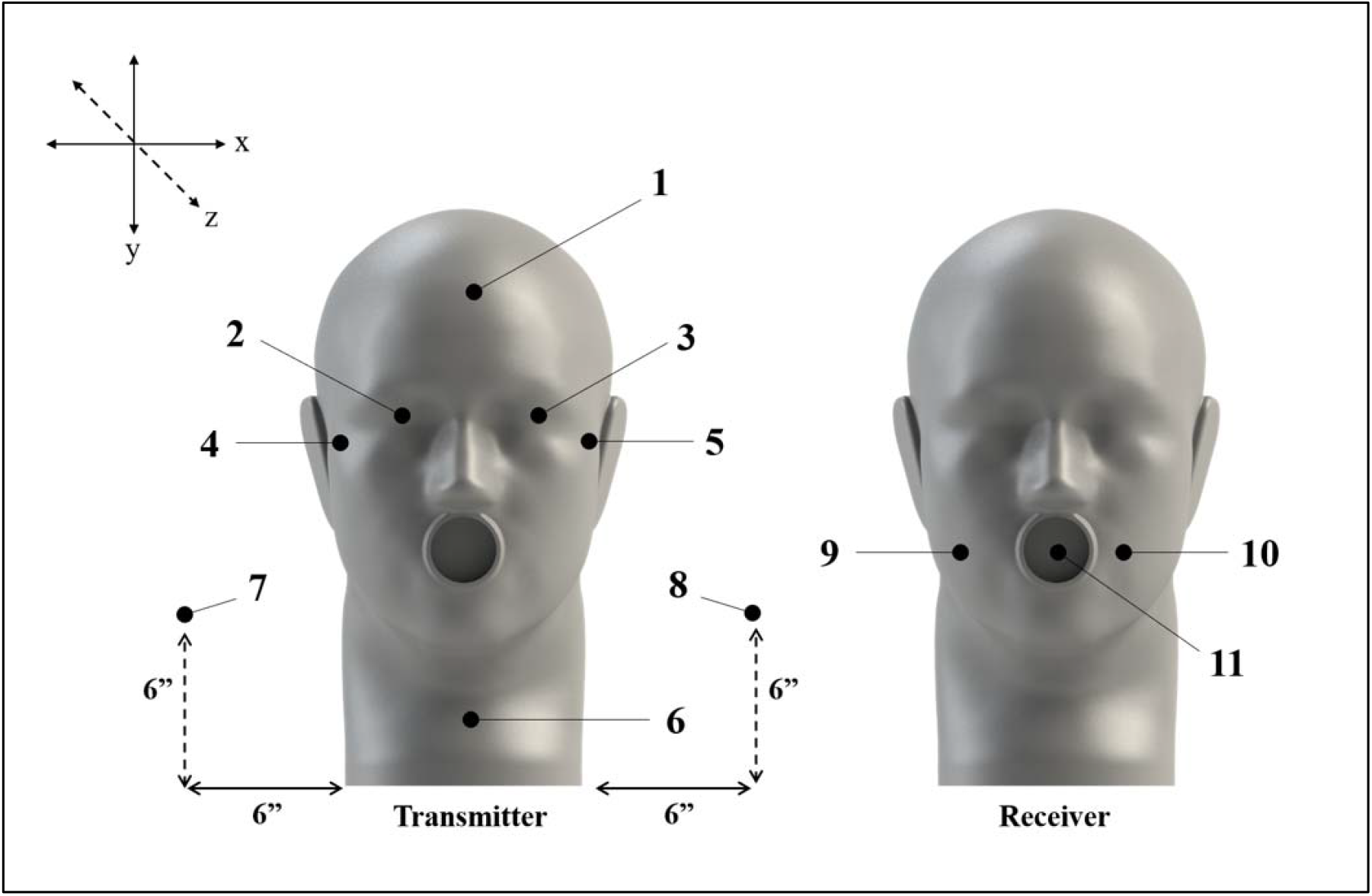
Headform Coupon Locations: 1) Forehead, 2) Left Eye, 3) Right Eye, 4) Left Cheek Bone, 5) Right Cheek Bone, 6) Neck, 7) Behind Left Shoulder, 8) Behind Right Shoulder, 9) Left Cheek, 10) Right Cheek, 11) Mouth.

A mucin solution (0.3%) was prepared using deionized ultra-filtered water and subsequently steam sterilized. The sterile mucin solution was cooled to room temperature and inoculated with a DNA marker at a concentration of ∼1 ng/mL and transferred into the pressure tank. Once the breathing machines reached their respective breathing rates, the valve on the pressure tank wa opened to initiate droplet production. Droplets were produced for 15 minutes, then the pressure tank valve was closed, and both breathing machines were turned off. Coupons were collected into sterile 1.5-mL microcentrifuge tubes using sterile forceps and subsequently extracted into 1 mL of IDTE (11-01-02-05; Integrated DNA Technologies). A 1-mL sample of residual mucin solution was also collected from the spray chamber. Coupon samples were vortexed vigorously for 5 seconds and mixed gently on the orbital rocker for 1 hour before being stored at 4 □.

### DNA Marker Analysis

The presence of λ-111 DNA was analyzed from 5 µL of each sample using a TaqMan-based absolute qPCR assay with a previously published primer-probe set.^16^ The presence of λ-111 DNA was analyzed from 5 µL of each sample using a TaqMan-based absolute qPCR assay with a previously published primer-probe set.^17^ Double-stranded genomic DNA from a different phage (PhiX174) was used as an exogenous positive control, also using a previously published primer-probe set.^17^ Absolute qPCR reactions were assembled using Brilliant III Ultra-Fast Probe qPCR Master Mix with low ROX (600890; Agilent, Santa Clara, CA) and were run on the Agilent AriaMX real-time PCR system. Three fluorescent dyes were monitored in multiplex reactions: 6-FAM-labelled λ DNA probe, Cy5-labelled PhiX174 DNA probe, and ROX as the passive reference dye. All reactions (singleplex and multiplex) were performed with 200-nM primer concentrations, 250-nM probe concentrations, and 10^2^ copies of PhiX174 DNA. A standard curve for absolute quantitation of λ-111 DNA was included in each 96-well plate and was constructed with serial dilutions of λ phage genomic DNA (10746782001; Sigma-Aldrich) ranging from 10^8^ to 10^0^ copies/µL. Two technical replicates were performed for each sample and duplicates with a variance greater than 1 C_q_ were excluded from further analysis and repeated until a lower variance was achieved.

A single qPCR reaction included either 5 µL of a sample or 1 µL of a standard, and all reactions included 1 µL of PhiX174 DNA at 10^2^ copies/µL to serve as the exogenous positive control. A negative control consisting of an unused coupon extracted into 1 mL IDTE was included in every experiment. A no-template control (NTC) assembled with IDTE was included in every 96-well plate. The quantity of λ-111 DNA copies was estimated for each reaction using AriaMX software. Reactions with C_q_ values greater than or equal to the negative control were assumed to have no λ-111 DNA. Quantity estimates less than 1 DNA copy were defined as below detection limit (BDL), which was 200 DNA copies/coupon.

### Data Analysis

The relative effectiveness of each PPE type was determined by calculating the percent reduction of DNA marker recovered from test coupons compared to positive control coupons collected in the same location. Percent reduction was calculated using Equation 1 followed by Equation 2. Samples that were below detection limit (BDL) were assigned half the detection limit (100 DNA copies).^18^ For statistical analysis, one-way ANOVA tests with Tukey’s post-test using non-log values were performed through Prism 9 (GraphPad; LaJolla, CA).

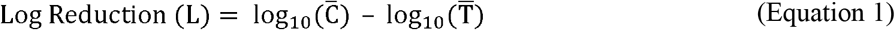

Where:

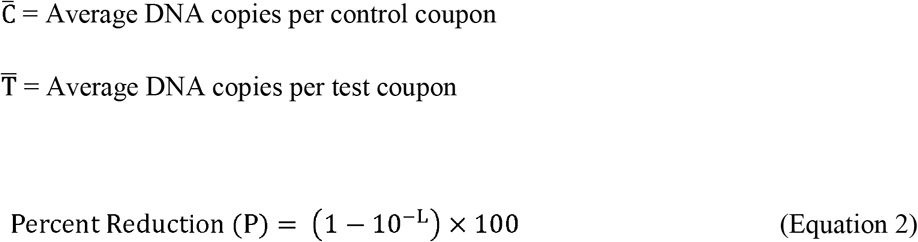

## Results

The droplet size across all three control runs ranged from 0.1 – 200 µm with a median diameter of ∼27 µm (Figure 3). Over 90% of droplets by volume ranged from 10 – 100 µm. The droplet mass output over the course of each run was estimated to be ∼1.2 g based on the volume output from the Spraytec droplet sizer data. The system only released droplets during exhalation and not during inhalation (data not shown). The mean DNA marker concentration sampled from the spray chamber was 6.46 ± 0.31 log_10_ DNA copies/µL across all tests performed. DNA marker was detected on 97% of all coupons evaluated, excluding negative control samples.

**Figure 3.**
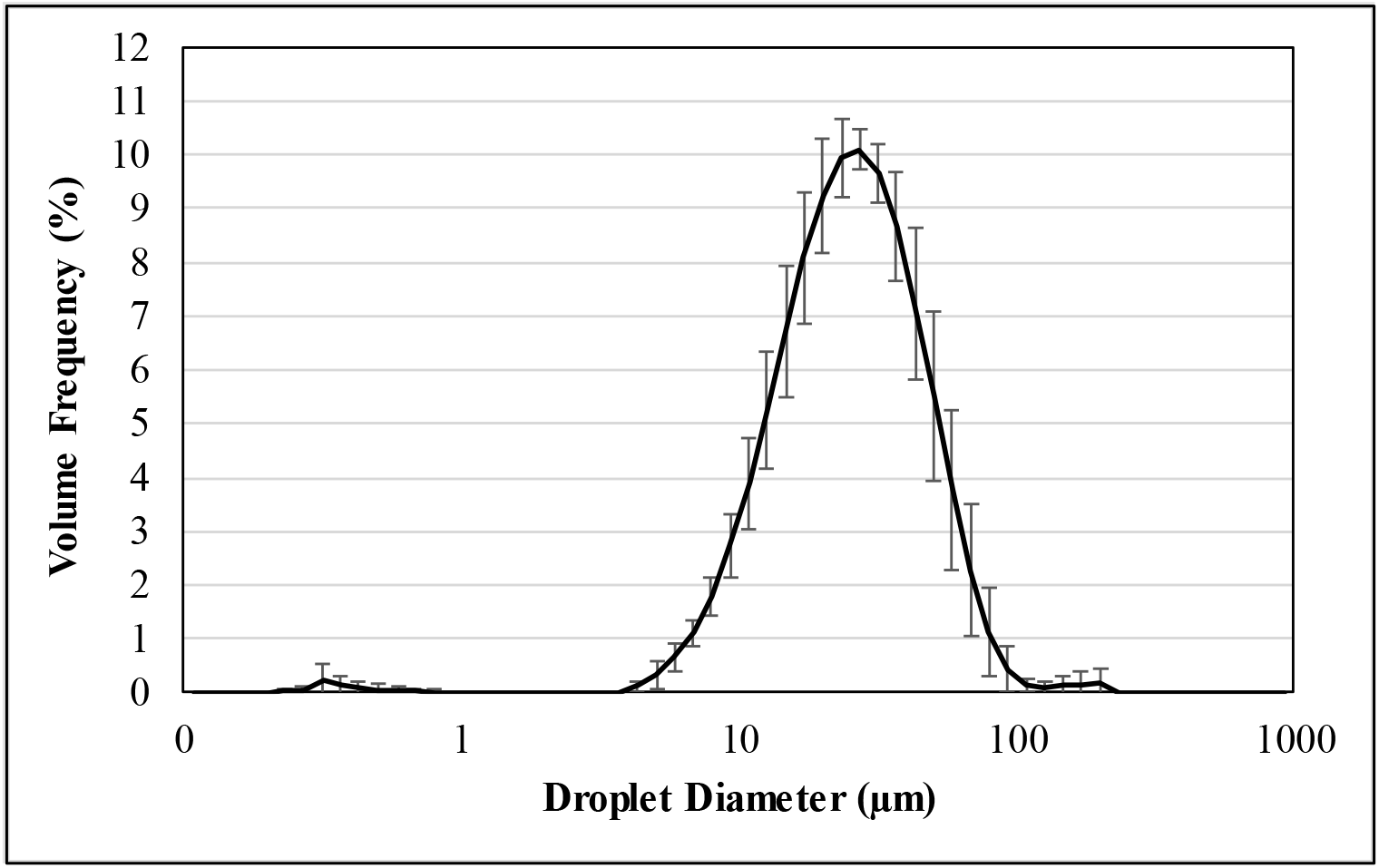
Mean Droplet Distribution for Triplicate Control Runs.

DNA was recovered from all coupons collected from the three control runs (no PPE), with the average number of DNA copies recovered per coupon being 5.17 ± 0.21 log_10_ DNA copies/coupon (Figure 4). The most contaminated control coupons were in the 1-ft position in front of the transmitter headform (5.42 ± 1.22 log_10_ DNA copies/coupon), followed by a decrease in contamination as the distance away from the transmitter increased up to 4 feet (4.87 ± 0.20 log_10_ DNA copies/coupon). DNA was not recovered from any of the negative control coupons.

**Figure 4.**
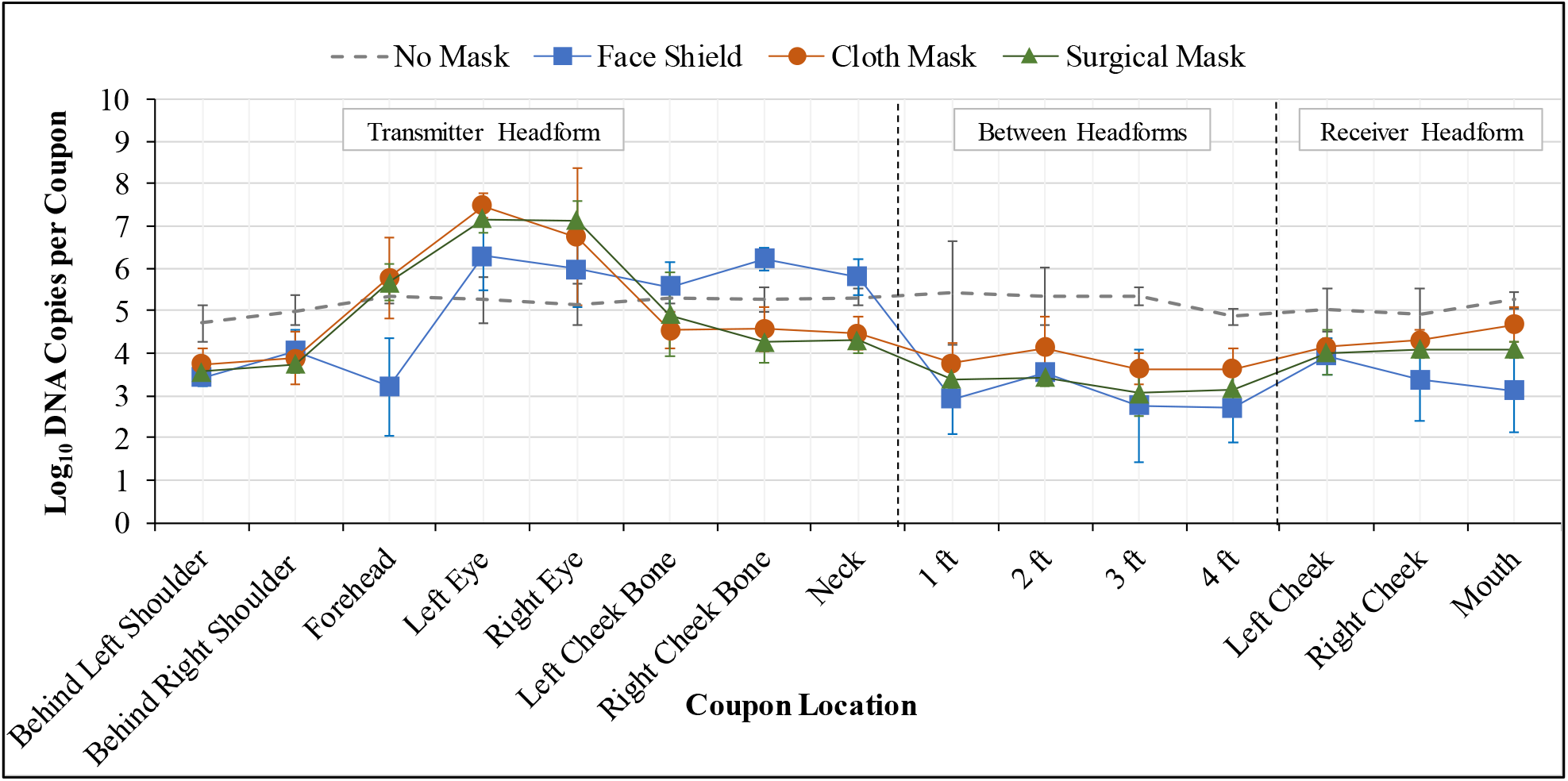
DNA Recovered from Sample Coupons after Droplet Transmission Tests.

All three types of PPE – face shield, cloth facemask, and surgical facemask – resulted in significant reductions of DNA contamination of the receiver headform (*p* < 0.0001, *p* = 0.0001, *p* = 0.0002, respectively) and between headforms (*p* = 0.04, *p* = 0.03, *p* = 0.03, respectively) (Table 1); no significant difference was found between PPE types. As distance from the transmitter increased, the significant difference also increased for all PPE types compared to the control.

**Table 1.** Percent Reduction of Recovered DNA Compared to Positive Control.

| System Location | Coupon Location | Cloth Facemask |  | Surgical Facemask |  | Face Shield |  |
| --- | --- | --- | --- | --- | --- | --- | --- |
|  |  | % Reduction | P-value | % Reduction | P-value | % Reduction | P-value |
| Transmitter headform | Behind shoulders | 90.9% | 0.32 | 93.8% | 0.81 | 92.0% | 0.99 |
|  | Forehead | -161.8% | 0.39 | -108.3% | 0.96 | 99.3% | 0.99 |
|  | Eyes | -10360.8% | <0.0001 | -8690.0% | 0.007 | -765.8% | 0.34 |
|  | Cheek bones | 81.2% | 0.98 | 74.8% | 0.99 | -458.1% | 0.29 |
|  | Neck | 85.9% | 0.87 | 90.2% | 0.84 | -207.4% | 0.12 |
| Between headforms | 1 foot | 97.8% | 0.08 | 99.0% | 0.08 | 99.7% | 0.08 |
|  | 2 feet | 94.1% | 0.09 | 98.8% | 0.06 | 98.4% | 0.06 |
|  | 3 feet | 98.1% | 0.006 | 99.5% | 0.006 | 99.7% | 0.007 |
|  | 4 feet | 94.4% | 0.005 | 98.1% | 0.004 | 99.3% | 0.003 |
| Receiver headform | Cheeks | 80.9% | <0.0001 | 87.3% | <0.0001 | 94.5% | <0.0001 |
|  | Mouth | 73.5% | 0.047 | 93.4% | 0.008 | 99.3% | 0.006 |

An increase in contamination was observed on the transmitter headform for all three PPE types, but in varying locations. Both the cloth facemasks and surgical facemasks resulted in significantly higher contamination of the transmitter’s eye region relative to the control (*p* < 0.0001, *p* = 0.007, respectively) and relative to the face shield (*p* = 0.0001 and *p* = 0.001, respectively). Higher contamination of the eye region was also observed with the face shield when compared to the control, but was not statistically significant (*p* = 0.34). Higher contamination of the transmitter’s forehead was observed for both facemasks, but the face shield resulted in lower contamination compared to the control; no statistically significant differences observed.

Conversely, the face shield produced a 458% increase in contamination of the transmitter’s cheek bone areas compared to the control, while both facemasks provided an average reduction of 78.0%; no statistically significant differences observed relative to the control or between the PPE tested. The face shield resulted in a 207.4% increase in contamination of the transmitter’s neck compared to the control, while the cloth facemask and surgical facemask demonstrated an average reduction of 88.1% – no statistically significant differences observed relative to the control; however, the neck contamination from face shield use was significantly higher compared to both the cloth facemask (*p* = 0.04) and the surgical facemask (*p* = 0.04).

## Discussion

The primary goal of this study was to evaluate the effectiveness of PPE at mitigating the spread of respiratory droplets when simulating the CDC close contact condition: a minimum of 15 minutes of contact at less than 6 feet. All three types of PPE – face shield, cloth facemask, and surgical facemask – demonstrated similar reductions in droplet transmission to the receiver headform.

For control testing (no PPE), a baseline level of contamination was recovered across all samples, which can likely be attributed to deposition of both droplets and aerosol particles (dried droplets). The highest contamination observed from control samples occurred at the 1-ft position in front of the transmitter headform, which then decreased as the distance away from the transmitter increased to 4 feet. This reduction in contamination can likely be attributed to droplet deposition onto coupons with larger droplets settling closer to the transmitter headform. Interestingly, higher levels of contamination were observed on the receiver headform relative to the 4-ft coupons. It is unclear why this occurred, however the breathing of the receiver headform may have influenced air patterns and caused aerosol particles (dried droplets) to interact with the coupons on the receiver. Because the testing was performed in a sealed room without ventilation, this likely allowed larger aerosol particles to interact with the sampling coupons, resulting in a baseline of detectable contamination for all tests.

Although the facial protection evaluated as part of this study demonstrated significant reductions in droplet transmission relative to the control, some level of contamination was detected on 96.5% of coupon samples across all tests where PPE was used. Similar to the control testing, this baseline level of detectable marker can likely be attributed to deposition of large aerosol particles (not droplets) over the course of each 15-minute test, which aligns with previous findings that face shields reduce user exposure to droplets by 96%, but do not contain all aerosols expelled by the user.^7,8^ The data from this study suggests this conclusion can be extended to surgical and cloth masks, and likely any face covering that does not provide a seal around the nose and mouth to mitigate the release of a user’s deflected exhalation. Although all PPE tested released some level of aerosol particles, they appear to reduce overall particulate aerosol contamination compared to the controls. The impact this finding has on the COVID-19 pandemic is relevant as demonstrated by the CDC acknowledgement that airborne transmission can occur under special circumstances (enclosed spaces, inadequate ventilation), and the use of all PPE tested may not allow for adequate source control.^1^ This highlights a need to better understand SARS-CoV-2 transmission by aerosols in addition to droplets.

Another trend for all three face coverings evaluated in this study is contamination detected on the transmitter headform during PPE use. With the face shield, higher contamination of all exposed surfaces of the transmitter headform (i.e., eyes, cheek bones, neck) was observed compared to the control, except for the forehead which is covered by a tight-fitting foam piece of the face shield. For both facemasks, higher contamination of the forehead and eyes of the transmitter headform were observed relative to the control. These results indicate the face shield forced the exhalation downward, while the facemasks forced the exhalation upwards. Both observations raise concern for self-contamination of contagious individuals, where the user’s face, facial accessories (e.g., eyeglasses), and/or nearby surfaces may become a source of contact transmission depending on the air flow path of the user’s exhalation. The concept of deflected exhaled air flow is intuitive as observed by users who experience fogging of their glasses when wearing facemasks. Moreover, during normal use, users would be expected to move their heads up and down, which could enlarge the contaminated area, especially for face shield users. When properly donned, facemasks are expected to be less affected by movement due to their more tight-fitting nature to the user’s face. More research is needed to better characterize the effect of head movement/position on localized contamination resulting from use by contagious individuals.

All simulation studies have limitations based on the conditions and test parameters selected. Breathing patterns and droplets produced by individuals will vary dramatically from person to person in real-world settings. A manikin was used for PPE use in this study, where actual fit on humans may impact the amount of air leakage around the devices. The study was performed in a non-ventilated setting, which may have influenced transmission of particles and to a lesser degree, droplets. Samples were also taken directly in front of the transmitter headform only – additional research is needed to assess the spread of droplets at various angles. Larger sample sizes would likely improve the statistical robustness and help overcome the inherent variability of aerosol testing. An assessment of PPE on the receiver headform would also likely produce informative results for public health measures. For this study, the simulation of droplet transmission in a real-world scenario was balanced with designing a reproduceable method, enabling comparisons of various types of PPE.

The focus of this study was to evaluate the effectiveness of PPE at mitigating the spread of respiratory droplets when simulating the CDC close contact condition: a minimum of 15 minutes of contact at less than 6 feet. All PPE tested were shown to be similarly effective at reducing droplet transmission up to 4 feet directly in front of the source, but were also shown to cause self-contamination. Variation in the location and severity of self-contamination observed suggests devices with differences in fit and form will likely result in different areas of the face being contaminated. However, the data from this study supports significant facial contamination should be expected with any PPE that does not provide a tight seal around the mouth and nose of the user. These findings have considerable implications for infection control procedures, drawing attention to the user’s face and nearby surfaces as potential sources of contact transmission when using such devices.

## Data Availability

The data that support the findings of this study are available from the corresponding author, D. Harnish, upon reasonable request.

## Acknowledgements

We would like to thank the Occupational Safety and Health Administration (OSHA) for funding this research under Prime contract #1605DC□19□A□0010. We would also like to thank the Air Force Civil Engineering Center (AFCEC) for providing a breathing machine for the study. We want to thank Mr. Mike McDonald for assistance setting up the test system.

